# Unsuspected clonal spread of Methicillin-resistant *Staphylococcus aureus* causing bloodstream infections in hospitalized adults detected using whole genome sequencing

**DOI:** 10.1101/2021.12.23.21268338

**Authors:** Brooke M. Talbot, Natasia F. Jacko, Robert A. Petit, David A. Pegues, Margot J. Shumaker, Timothy D. Read, Michael Z. David

**Affiliations:** Graduate School of Biological and Biomedical Sciences, Emory University, Atlanta, Georgia, USA; Division of Infectious Diseases, Department of Medicine, University of Pennsylvania, Philadelphia, PA, USA; Division of Infectious Diseases, Emory University School of Medicine, Atlanta, Georgia, USA; Division of Infectious Diseases, Department of Medicine, Perelman School of Medicine at the University of Pennsylvania, Philadelphia, PA, USA

**Keywords:** Staphylococcus aureus, bloodstream infections, hospital epidemiology, outbreak detection, infection prevention

## Abstract

**Background:** Though detection of transmission clusters of methicillin-resistant *Staphylococcus aureus* (MRSA) infections is a priority for infection control personnel in hospitals, the transmission dynamics of MRSA among hospitalized patients with bloodstream infections (BSIs) has not been thoroughly studied. Whole genome sequencing (WGS) of MRSA isolates for surveillance is valuable for detecting outbreaks in hospitals, but the bioinformatic approaches used are diverse and difficult to compare.

**Methods:** We combined short-read WGS with genotypic, phenotypic, and epidemiological characteristics of 106 MRSA BSI isolates collected for routine microbiological diagnosis from inpatients in two hospitals over 12 months. Clinical data and hospitalization history were abstracted from electronic medical records. We compared three genome sequence alignment strategies to assess similarity in cluster ascertainment. We conducted logistic regression to measure the probability of predicting prior hospital overlap between clustered patient isolates by the genetic distance of their isolates.

**Results:** While the three alignment approaches detected similar results, they showed some variation. A pangenome-based alignment method was most consistent across MRSA clonal complexes. We identified nine unique clusters of closely-related BSI isolates. Most BSI were healthcare-associated and community-onset. Our logistic model showed that with 13 single nucleotide polymorphisms the likelihood that any two patients in a cluster overlapped in a hospital was 50 percent.

**Conclusions:** Multiple clusters of closely related MRSA isolates can be identified using WGS among strains cultured from BSI in two hospitals. Genomic clustering of these infections suggest that transmission resulted from a mix of community spread and healthcare exposures long before BSI diagnosis.

**Summary:** Multiple clusters of closely related MRSA bloodstream infections were identified using WGS in two hospitals using three bioinformatic workflows. Genomic epidemiology suggests that transmission resulted from a mix of community spread and healthcare exposures long before symptom onset.

## Introduction

*Staphylococcus aureu*s infections are a common cause of morbidity and mortality, ranging from skin and soft tissue infections to severe, invasive disease such as pneumonia, osteomyelitis, and bloodstream infections (BSI). *S. aureus* caused nearly 119,000 BSIs and 20,000 associated deaths in 2019 [1]. These infections are exacerbated by the emergence of methicillin-resistant *S. aureus* (MRSA) strains which are resistant to treatment with conventional ß-lactam antibiotics. Concerted national infection control efforts have decreased MRSA healthcare-associated infections (HAIs) in the United States (U.S.), particularly BSIs caused by MRSA strains historically associated with HAIs. However, the decrease in MRSA BSIs in the U.S. has slowed since 2013, and community-onset infections have recently made up the largest proportion of cases [1].

Asymptomatic *S. aureus* carriage is a risk factor for infection, and in addition to the well-characterized reservoir of nasal carriage [2], the intestinal tract, pharynx, skin, and perineal areas are also important reservoirs [3]. Colonization makes elimination of this pathogen especially difficult, as detecting carriage or transmission from person to person can occur long after the initial exposure. Onset of a state of disease (i.e., clinically significant infection) is influenced by bacterial virulence, human host factors, and triggers such as skin trauma or underlying illnesses that predispose to opportunistic infections [4]. Spread of *S. aureus* from direct or indirect contact with colonized individuals, contamination of an intermediate third person (such as a healthcare worker [5]), or through environmental reservoirs has resulted in numerous outbreaks in hospitals [6] and community settings [7–9]. Importantly, though detection of transmission clusters of MRSA infections is a priority of infection control personnel in hospital settings, the transmission dynamics of MRSA among hospitalized patients with BSIs has not been thoroughly studied.

Whole genome sequencing (WGS) of bacterial genomes provides high resolution to identify clusters of closely related MRSA isolates, from which recent transmission may be inferred. Because of this, WGS is an increasingly important tool used for infectious disease surveillance. This is possible with improved access and ease of use of open-source bioinformatic resources, lower costs associated with implementation in smaller lab or clinical lab settings, and the expansion of available sequence data in public databases [10,11]. Investigators now have many choices of methods that can be used to create WGS cluster detection workflows. The choice of genome alignment method is particularly important for comparative genomics because it is the foundation for matching gene homology. Various alignment strategies include reference-free or reference-dependent methods, and each have their own trade-offs for sensitivity, specificity, and the amount of genetic information used in a final comparison of sequences [12].

To date, epidemiological investigations use genetic thresholds for ease of decision making to determine the relatedness among *S. aureus* isolates, which either identifies clusters of related infections or rules out cases from an investigation [6,13,14]. One simple threshold is to quantify single-nucleotide polymorphisms (SNPs) differing between genome sequences. Comparison of these differences through multisequence alignment and phylogenetic reconstruction is used as a means of estimating likelihood of a recent common ancestor and thus suggesting a transmission event [15,16]. Choice of a reference sequence, existing genetic diversity among samples, sample size, and bioinformatic tools and workflows can all impact which and how many SNPs are detected in a single set of samples. It is necessary to explore the consistency of SNP identification when different alignment tools are used.

To learn more about potential risk factors for transmission of MRSA BSI in hospitalized patients, we report a retrospective analysis of MRSA BSI at two hospitals in a single university system during a 12-month period. We compared core genome sequences from the isolates to detect putative transmission events occurring between BSI patients. We then examined epidemiological data and genetic traits of isolates shared within and between patients in each cluster. We also tested the consistency of observed genetic differences between isolates using different genome sequencing alignment tools used to identify SNP differences.

## Methods

### Patient cohort selection

We identified all patients diagnosed with a MRSA BSI between July 2018 and June 2019, admitted to either of two hospitals of the University of Pennsylvania hospital system. The Hospital of the University of Pennsylvania (HUP) is a 625-bed academic tertiary and quaternary care medical center in West Philadelphia with approximately 32,000 patient admissions, 633,000 outpatient visits, and 40,000 Emergency Department visits annually. The Penn Presbyterian Medical Center (PMC) is a 324-bed urban community hospital in West Philadelphia with 12,000 admissions, 130,000 outpatient visits, and 26,000 Emergency Department visits annually. A single case of MRSA BSI was defined as any instance of a MRSA isolate collected from blood of any patient at HUP or PMC during the study period. Each subject was only included once. The study was approved by the University of Pennsylvania IRB and given a waiver of consent, as the study was retrospective and no data or samples were collected specifically for research purposes.

### Isolate selection and DNA sequencing

Isolates were obtained from a biobank that collected all clinical MRSA isolates cultured from routine diagnostic studies in the HUP Clinical Microbiology Laboratory during the study period. They included all sequential MRSA BSI isolates from the study population. Isolates were screened for phenotypic resistance to twenty antibiotics using the Vitek 2 automated system, and susceptibility and resistance were assigned in accordance with CLSI protocols [17]. A 1μL loopful of each frozen isolate was streaked onto blood agar and incubated overnight at 37°C. A single isolated colony was selected from each plate and grown again on blood agar overnight at 37°C. A 10μL loopful of each isolate was then frozen in a bead beating tube and underwent whole genome sequencing (WGS) at the Penn/Children’s Hospital of Philadelphia Microbiome Center. Sequencing libraries were prepared using the Illumina Nextera library preparation kit, and sequenced on the Illumina MiSeq. Sequences were made publicly available through the NCBI Sequence Read Archive (Bioproject PRJNA751847).

### Bioinformatic pipelines

Paired-end 150 bp FASTQ files were passed through the “Bactopia” workflow to assess data quality, assemble contigs, call MLST, SCC*mec* type, antibiotic resistance and virulence genes [18]. To compare SNP-based core-genome multiple-sequence alignments, the total number of assembled contigs or subsets grouped by clonal complex (CC) were passed through two reference-based core SNP alignment tools, Snippy (v.4.6.0) [19] and Parsnp (v.1.5.6) [20], or concatenated and aligned using tools within the PIRATE workflow [21]. ClonalFrameML [22] and maskrc-svg (v0.5)(https://github.com/kwongj/maskrc-svg) were used to identify and mask possible recombinant regions with in the core-genome alignment. Pairwise SNP distances for each alignment were calculated using snp-dists [23]. Maximum likelihood trees were created using IQ-Tree (v2.1.2) [24] and visualized using the ggTree package [25]. Bootstrap values of the most recent common ancestral node were calculated for 1000 repetitions. We used N315 strain genome (GCF_000009645.1) for total isolate alignments and CC5-specific alignments; for the CC8 specific alignment we used NCTC 8325 as reference (GCF_000013425.1). Phylogenetic similarity across alignment strategies was measured by calculating cophenetic correlation [26] between SNP distance matrices and estimated phylogeny tip distance and Robinson-Foulds distances [27] between trees generated with the different alignments as well as randomly generated trees using ape version 5.5 [28]. The pangenome-mapping of the 104 isolates was used because the tree had the strongest correlation between the estimated phylogeny, high tree structure similarity to other alignment tools, and conservation of SNP distances between alignment strategies (Table 1; Fig 1A-B).

**Table 1.** Comparability phylogenetic fit of alignment methods using Cophenetic correlation (R^2^), alignment size, and Robinson-Foulds (RF) comparison^a^ by alignment method

| Method | Total isolates (N=104) |  |  | CC5-specific isolates (N =40) |  |  | CC8-specific isolates (N = 55) |  |  |
| --- | --- | --- | --- | --- | --- | --- | --- | --- | --- |
| | $R^2$ | Alignment Size (bp) | RF-values | $R^2$ | Alignment Size (bp) | RF-value | $R^2$ | Alignment Size (bp) | RF-values |
| **Pangenome-pipeline** | 0.984 | 2,141,357 | 56,52,200 | 0.984 | 2,182,742 | 10,8,72 | 0.987 | 2,176,046 | 40,36,104 |
| **Pseudo-read-pipeline** | 0.983 | 2,839,469 | 46,56,202 | 0.993 | 2,839,469 | 10,10,72 | 0.999 | 2,821,361 | 34,40,104 |
| **Assembly-pipeline** | 0.929 | 2,163,693 | 52,46,202 | 0.995 | 2,497,454 | 8,10,72 | 0.999 | 2,482,874 | 34,36,104 |

<sup>a</sup>Row alignment method compared to each other alignment method and a random tree of the same number of phylogenetic tips

**Figure 1.**
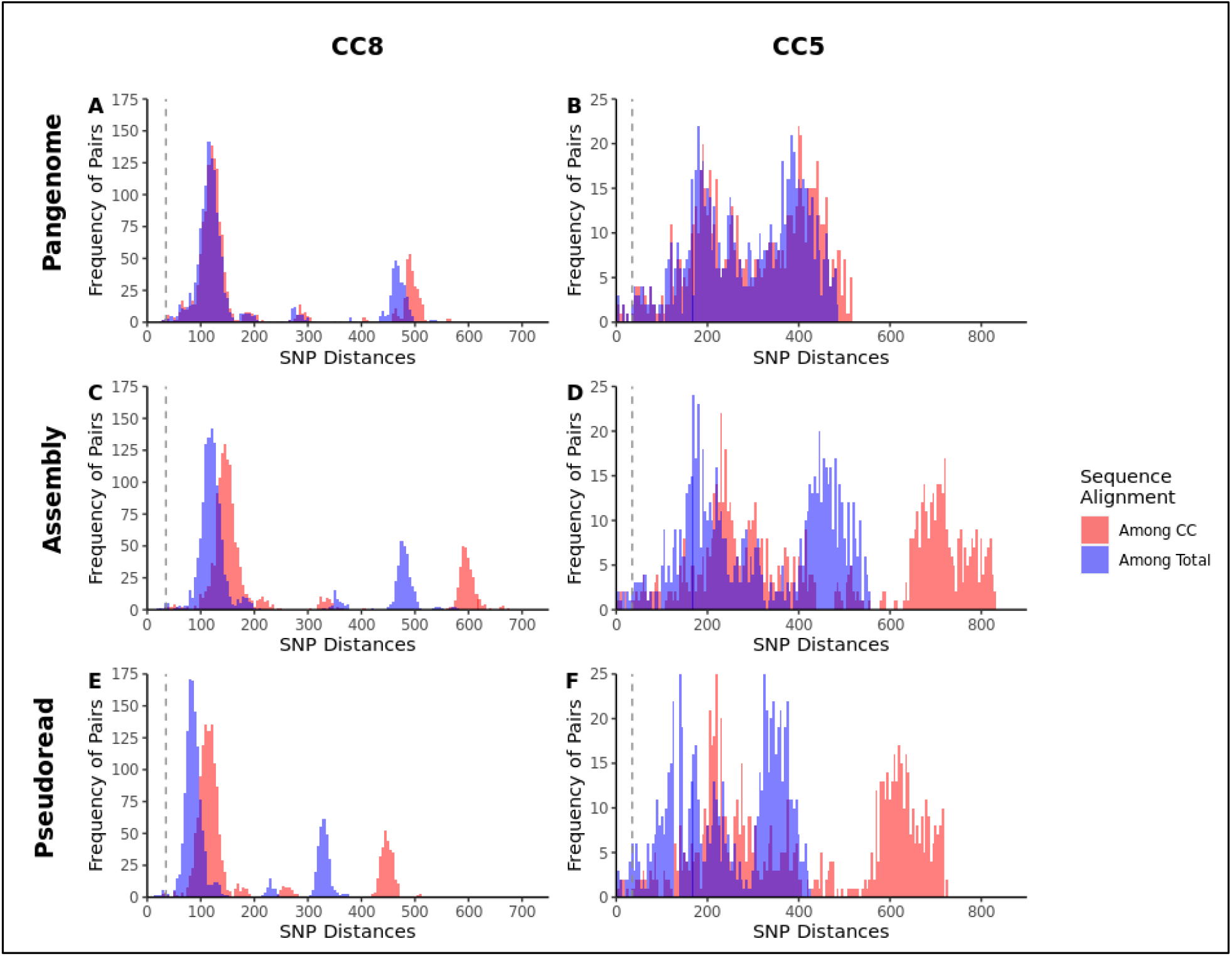
Frequencies and distribution of single nucleotide polymorphism distances between isolates varies by alignment tool. Whole-genome sequence alignments were created from MRSA alignments using either all 104 isolate sequences (blue) or isolates categorized only as clonal complex (CC) 8 and 5 (red), and using three alignment strategies: Pangenome-pipeline (A,B), Assembly-pipeline (C,D), and Pseudoread-pipeline (E,F). Pairwise single nucleotide polymorphism (SNP) distances were plotted for each alignment strategy.

### Epidemiological Investigation of Clustered Isolates

We analyzed core genomes with pairwise SNP differences less than or equal to two thresholds to detect patient-patient transmission: 35 SNPs and 15 SNPs. The threshold of 35 SNPs was based on the approximate cutoff found in within-patient versus between-patient BSI MRSA lineages in a hospital setting [6,29]. The more stringent threshold of 15 SNPs for core genome comparisons was proposed as a conservative cutoff for recent inter-patient MRSA transmission [13]. Demographic data, comorbidities, Pitt Bacteremia Score, source of BSI, and in-patient mortality were collected for each study subject by abstraction from the electronic medical record (EMR). A BSI was considered to be healthcare-associated (HA) if the index blood culture was drawn >48 hours after hospital admission; healthcare-associated, community-onset (HACO) if the index culture was obtained <48 hours after admission or in the community setting, and if the subject had one or more previous healthcare risk factors (hospitalization, surgery, hemodialysis, or stay in a nursing home or another residential medical facility in the previous year; or presence of an indwelling intravascular catheter at time of culture); and community-associated (CA) if the index culture was obtained <48 hours after admission or in the community setting and the subject lacked these healthcare risk factors. Demographic and clinical characteristics were tabulated for all subjects. The EMR was examined for evidence of overlap or sequential hospital/unit stays among cluster subjects, defined as cases in which subjects’ MRSA isolates from >1 subject clustered at the 35 SNP threshold, and visualized using the vistime package (https://github.com/shosaco/vistime). Dates of admission and discharge for all hospital stays at any of the four networked hospitals in the one year before the first collected BSI isolate in a cluster and one year after the last collected BSI isolate in the cluster were recorded for each cluster subject. In addition to HUP and PMC, any relevant hospitalizations were recorded at Pennsylvania Hospital (PH) and at a single, University of Pennsylvania long-term acute care hospital in Philadelphia during the year prior to their index BSI culture. PH is a 481-bed urban community hospital located in the Society Hill district of Philadelphia with over 27,000 hospital admissions, over 24,000 Emergency Department visits, and 201,000 outpatient visits annually.

Logistic regression modelling was used to assess the predictive power of pairwise SNP distances and likelihood of patient temporal hospitalization overlaps. Goodness of fit was assessed using a receiver operating characteristic (ROC) curve and measuring the area under the curve. All analyses were conducted in R studio (v1.4.1106)[30] run with R version 4.0.4, and final figures labelled in InkScape (v0.92.5) [31]. Analysis code is available at https://github.com/Read-Lab-Confederation/MRSA_bloodstream_clusters.

## Results

### Distribution of patient demographics and isolate characteristics

We screened all patients diagnosed with a MRSA BSI at three academic hospitals between July 2018 and June 2019, identifying 106 qualifying subjects (see Materials and Methods). Of the BSI source sites that could be identified from EMR, skin site infections made up 19% and central venous catheter infections made up 14% (Table 2). Among included subjects, 16% died while in the hospital.

**Table 2.** Demographics and clinical outcomes of subjects with MRSA bloodstream infection (n=106)

| Demographic Characteristic | Number (%) Patients | Clinical Characteristic | Number (%) Patients |
| --- | --- | --- | --- |
| **Total** | 106 | **Total** | 106 |
| **Age Group** |  | **Source of BSI** |  |
| 20-29 | 12 (11%) | Arteriovenous graft | 4 (4%) |
| 30-39 | 13 (12%) | Central venous catheter infection | 15 (14%) |
| 40-49 | 16 (15%) | Device infection | 4 (4%) |
| 50-59 | 18 (17%) | Respiratory source | 2 (2%) |
| 60-69 | 32 (30%) | Skin site | 20 (19%) |
| 70+ | 15 (14%) | Surgical site | 4 (4%) |
| **Sex** |  | Other | 3 (3%) |
| Female | 51 (48%) | Unknown | 52 (49%) |
| Male | 55 (52%) | **Hospital of BSI diagnosis** |  |
| **Race** |  | Hospital A | 65 (61%) |
| Asian | 1 (1%) | Hospital B | 41 (39%) |
| White | 50 (48%) | **Infection setting** |  |
| Black | 49 (46%) | HA | 22 (21%) |
| Other/Unknown | 6 (6%) | CA | 11 (10%) |
| **Ethnicity** |  | HACO | 71 (68%) |
| Hispanic/Latino | 2 (2%) | **In-hospital death<sup>a</sup>** |  |
| Non-Hispanic/Latino | 99 (93%) | No | 88 (83%) |
| Unknown | 5 (5%) | Yes | 18 (17%) |
|  |  | **Pitt bacteremia Score** |  |
|  |  | Mean (SD) | 2.1 (2.6) |
|  |  | Median (Range) | 1.00 (0, 10.0) |

<sup>a</sup>Indicates death prior to discharge during the index MRSA BSI hospitalization. Abbreviations: BSI: bloodstream infection; CA: community-associated; HA: healthcare-associated; HACO: healthcare-associated, community-onset; SD: standard deviation.

From each individual, single MRSA samples were isolated and sequenced, of which 105 had sufficient coverage for further analysis and 104 isolates were determined to be *Staphylococcus aureus*. One isolate was identified by WGS as *Staphylococcus argenteus* and was excluded from further analysis. Among the 104 genomes, 55 were assigned to CC8, 49 of which were USA300 strains; 40 were assigned to CC5; and the remaining nine isolates were in CC30, CC72, and CC78.

### Assessment of sequence alignment strategies

We generated multiple alignments of the MRSA isolates using three approaches to determine their effect on pairwise SNP distances between isolates (Fig. 1). The three approaches were (1) randomly fragmenting assembled genomes to create “pseudoreads” and mapping these to a reference genome using Snippy (“pseudoread-pipeline”); (2) mapping assembled genomes to the reference using Parsnp (“assembly-pipeline”); and (3) aligning concatenated core genes that were identified using the PIRATE tool (“pangenome-pipeline”). Alignments generated with all 104 isolates included had lower pairwise distances compared to alignments generated only with isolates of the same CC. This was because the alignments with larger numbers of more diverse strains always covered a smaller amount of the chromosome (Table 2). Pairwise SNP distances produced by the pangenome-mapping method were consistent between the CC groups and whole species alignments (Fig 1A-B), whereas the pairwise SNP distances produced by pseudoread-mapping and assembly-mapping methods were greater when smaller groups of more closely-related strains were the input (Fig. 1 C-F).

### Suspected transmission clusters

Using alignments containing all 104 isolates for each pipeline, 29 isolates grouped into nine clusters (C1-C9) where isolates had less than a 35-SNP threshold difference from at least one other subject isolate (Table 3). The psuedoread-pipeline clustered 29 isolates, the assembly-pipeline clustered 21, and the pangenome-pipeline clustered 19. Five clusters contained isolates from CC5, three clusters were CC8, and one cluster was CC30. Three clusters also contained sub-cluster groups of isolates which fell within the SNP threshold for only some of the tools. One cluster (C7) was only detected by assembly-mapping. The median cluster size was three isolates (range 2 - 6). Blood culture collection date differences between clustering isolates was wide, with the longest period being 265 days (C1), and the shortest being 12 days (C6). Median SNP differences were diverse across clusters, and smaller differences did not correlate with shorter collection date differences.

**Table 3.** Summary of suspected MRSA transmission clusters identified through Psuedoread-, Assembly-, and Pangenome-alignment pipelines among 104 sequential MRSA bloodstream infection patients at 2 hospitals

| Cluster | MRSA Isolate | Clonal Cluster | Number of isolates | Median Pairwise SNP Difference (Range) |  |  | Median Collection Date Difference, Days (Range) |
| --- | --- | --- | --- | --- | --- | --- | --- |
|  |  |  |  | Pseudoread-pipeline | Assembly-pipeline | Pangenome-pipeline |  |
| C1 | SAMN20960259,<br>SAMN20960281,<br>SAMN20960331 | CC5 | 3 | 11 (3-12) | 16 (4-16) | 14 (5-16) | 177 (110 - 265) |
| C2 | SAMN20960260,<br>SAMN20960274 | CC5 | 2 | 6 | 7 | 7 | 61 |
| C3a | SAMN20960263,<br>SAMN20960326,<br>SAMN20960280,<br>SAMN20960314,<br>SAMN20960328 | CC5 | 5 | 35 (20 - 46) | 44 (26-62) | 50 (36 - 62)<sup>a</sup> | 128 (7 - 241) |
| C3b | SAMN20960280,<br>SAMN20960314,<br>SAMN20960328 | CC5 | 3 | 25 (20 - 25) | 32 (26-36) | 39 (36 - 39)<sup>a</sup> | 113 (37 - 150) |
| C4 | SAMN20960270,<br>SAMN20960325 | CC5 | 2 | 20 | 26 | 24 | 189 |
| C5a | SAMN20960271,<br>SAMN20960343 | CC5 | 4 | 29 (6 - 35) | 44 (10-53)<sup>a</sup> | 41 (33 - 46)<sup>a</sup> | 119 (56 - 237) |
| C5b | SAMN20960271,<br>SAMN20960343 | CC5 | 2 | 29 | 42<sup>a</sup> | 33 | 237 |
| C5c | SAMN20960298,<br>SAMN20960324 | CC5 | 2 | 6 | 10 | 7 | 78 |
| C6a | SAMN20960276,<br>SAMN20960282,<br>SAMN20960287,<br>SAMN20960293,<br>SAMN20960301,<br>SAMN20960306 | CC8 | 6 | 29 (15 - 42) | 39 (21-62) | 38 (26 - 53)<sup>a</sup> | 54 (12 - 121) |
| C6b | SAMN20960276,<br>SAMN20960282,<br>SAMN20960293,<br>SAMN20960301,<br>SAMN20960306 | CC8 | 5 | 26 (15 - 31) | 35 (21-40) | 37 (26 - 43) | 66 (12 - 121) |
| C6c | SAMN20960276,<br>SAMN20960282,<br>SAMN20960293,<br>SAMN20960306 | CC8 | 4 | 23 (15 - 30) | 32 (21 - 36) | 34 (26-38) | 63 (32 - 121) |
| C7 | SAMN20960299,<br>SAMN20960305,<br>SAMN20960334 | CC8 | 3 | 34 (30 -34) | 39 (36 - 41) | 45 (41 - 50) | 80 (24 -104) |
| C8 | SAMN20960313,<br>SAMN20960323 | CC8 | 2 | 1 | 1 | 1 | 28 |
| C9 | SAMN20960316,<br>SAMN20960337 | CC30 | 2 | 23 | 25 | 22 | 67 |

<sup>a</sup>Partial or no detection of isolates as part of the cluster

### Phylogenetic analysis of patient isolates

BSI isolates, and those isolates falling into possible transmission clusters, spanned across significantly divergent clades of CCs (SH-aRLT and uBS values >70) (Fig 2A). Candidate transmission clusters formed distinct sub lineages within the clades. The largest cluster, C5, diverged significantly from other CC8 isolates and were identified as part of the CC8c lineages, a strain of USA500 as defined by the phylogenetic subtyping scheme by Bowers et al. [32]. Cluster and non-cluster isolates had a similar distribution for infection setting, with most BSI isolates categorized as HACO (68%). At a 15-SNP threshold, only isolates in clusters C1, C2, C5(a,c), and C8 remained clustered. All isolates were susceptible to vancomycin and daptomycin, but isolates in both the CC5 and CC8 clades contained multi-antibiotic resistant phenotypes to beta-lactams (as expected for MRSA) and quinolones. Together, these data suggested that multiple lineages of MRSA associated with BSI had the potential to transmit between patients, and those clusters had a higher likelihood of harboring multi-class resistance phenotypes.

**Figure 2.**
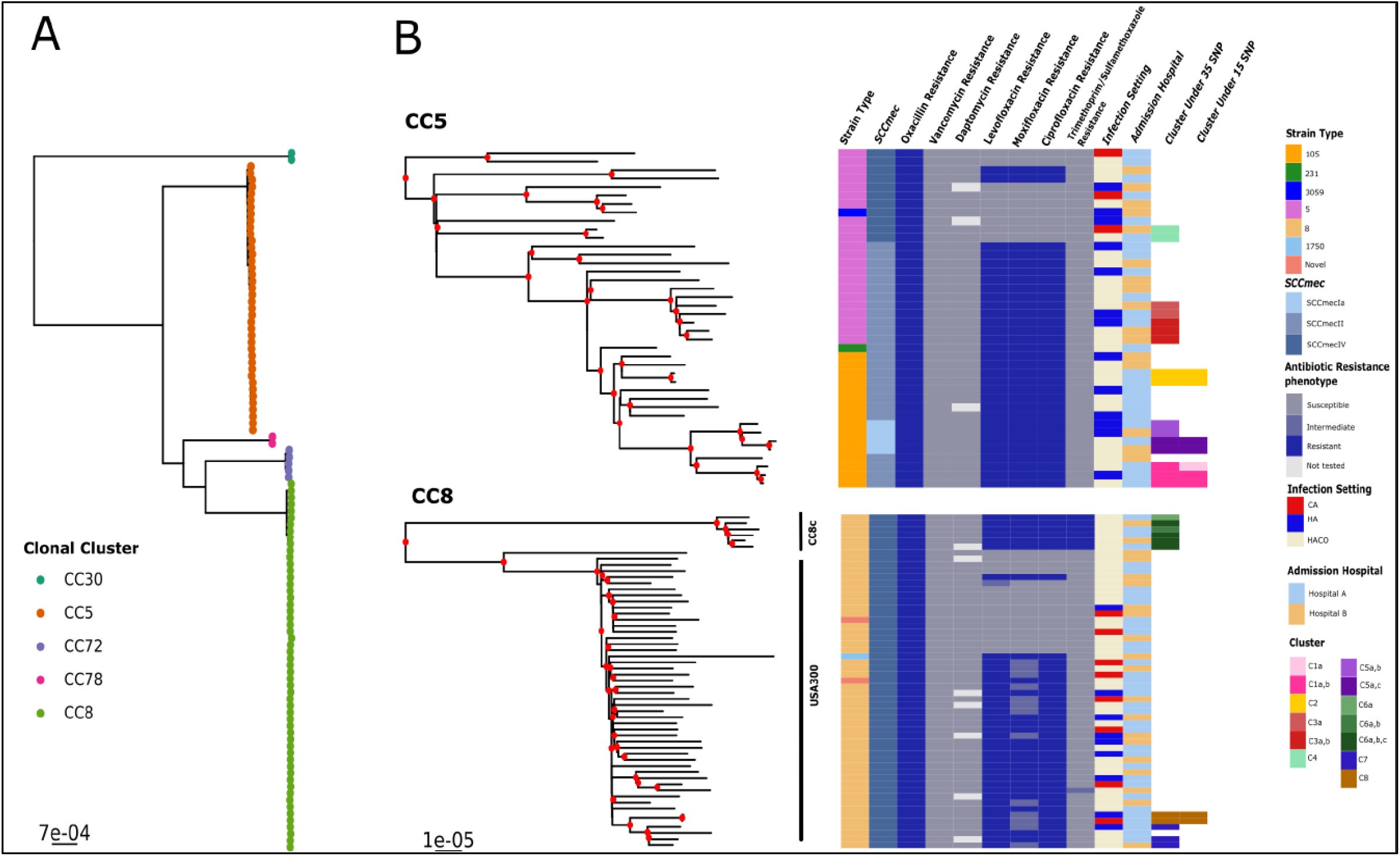
Suspected transmission clusters fall into distinct clonal groups. Maximum likelihood trees were generated from the PIRATE alignment of 104 isolates and visualized using ggtree. (A) Tree indicating clades containing individual clonal complexes (CCs). (B) Subtrees from the complete maximum likelihood trees for the two most abundant CCs. Nodes with bootstrap values >= 70 are marked in red. Heat maps show strain type, SCCmec element type, and resistance phenotype for indicated antibiotics per sequence, infection setting (Healthcare-associated (HA), Community associated (CA) and Healthcare-associated community-onset (HACO)), admission hospital, and transmission cluster at a threshold of 35 SNPs or 15 SNPs.

### Genomic similarity predicts overlapping hospital stay in transmission clusters

To compare common healthcare exposures among clustered isolates, we examined the hospitalization history for every subject within a cluster at four networked hospitals in the University of Pennsylvania system one year before the first index BSI isolate and one year after the last patient index isolate. Six had subjects with overlapping hospital stays (Fig. 3), of which three had median SNP distances between 1-16 core SNPs with corresponding hospital unit overlaps (Table 3; Fig. 3). Cluster C5c had a median SNP difference of seven (range 6-10 SNPs across tools) with no common hospital overlap. In comparison, cluster C4 also had no subjects with overlapping hospital admissions prior to their index BSI cultures, but the median SNP distance range of 20 - 26 SNPs across tools.

**Figure 3.**
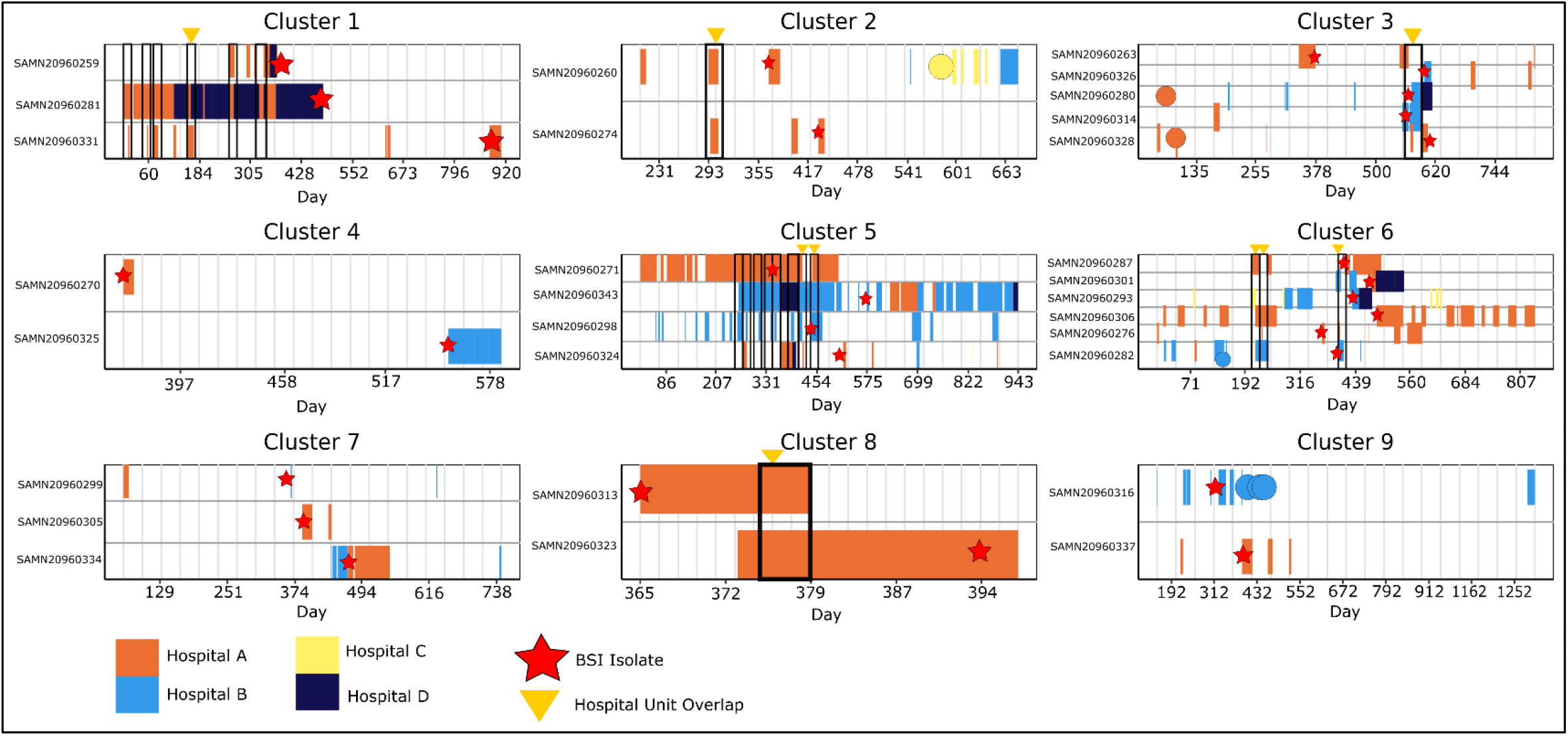
Hospitalization history among patients in genomic BSI clusters. Hospitalization history at 4 study hospitals (A, B, C, and D) up to 365 days before the date of the earliest MRSA bloodstream isolate culture in each cluster (relative Day 0) and up to 365 days after the latest MRSA bloodstream isolate in the cluster. Note that bloodstream infections were only included at hospitals A and B. Rows represent the hospitalization history of each patient associated with a sequenced cluster isolate. Colored rectangles and circular marks represent individual hospitalization durations (rectangles) or one-day admissions (circles); the color indicates Hospital, A, B, C, or D. Black outlined boxes represent areas where two or more patients overlapped in the same hospital at the same time. Red stars indicate the date of collection of the sequenced BSI isolate for each patient. Yellow triangles indicate a hospitalization where two or more patients overlapped in the same hospital unit.

The SNP threshold is a critical component for identifying clusters for further investigation. We therefore performed a logistic regression to determine if there was an association between likely hospital exposure and SNP threshold (Fig. 4). The log odds of clustered patient pairs overlapping in the same hospital decreases by 0.065 with every increase of one SNP (p=0.05). Our logistic model showed that with 13 SNPs the likelihood that any two patients in a cluster overlapped in a hospital was 50 percent (Fig. 3A). Pairwise SNP distances that were greater than 13 SNPs trended toward patients with no overlapping hospital stays in the year prior to their MRSA bacteremia infection. The area under the curve of the receiver operating characteristic analysis indicates that we could classify true positive exposures (i.e., known prior overlapping hospitalizations) 66% of the time from the SNP threshold (Fig. 3B).

**Figure 4.**
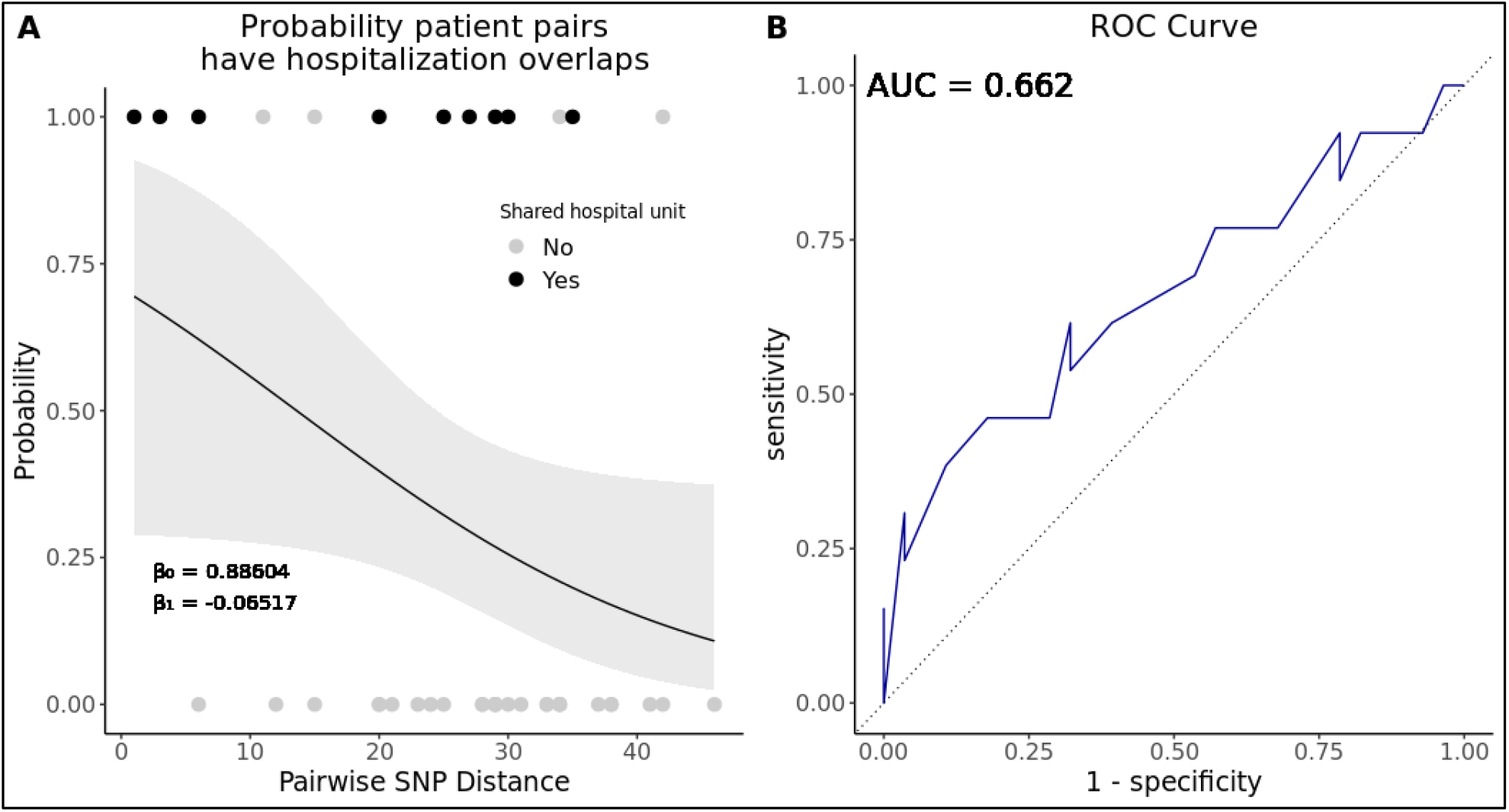
Higher SNP distances trend toward ruling out hospital overlaps between clustering patients. (A) Logistic regression model indicating the relationship between patient pairs overlapping in the same hospital at the same time (prior to the diagnosis of an index MRSA bloodstream infection) and the pairwise SNP distance. Points indicate the true result for each pair as overlapping (1.0) or not overlapping (0). The color of the points indicates whether hospital overlap patient pairs also overlapped (black) or did not overlap (gray) in the same hospital unit. Gray ribbon indicates the 95% confidence interval. (B) Receiving operating characteristic (ROC) curve of the logistic model in A. Area under the curve (AUC) = 0.662.

## Discussion

In our investigation we combined clinical and bacterial genome data to describe a large cohort of US MRSA BSI patients. The predominant genetic backgrounds of MRSA isolates in this study were consistent with reports documenting the expansion of CC8 and CC5 MRSA strains in the US, which have been documented to cause both healthcare- and community-associated infections [33]. Therefore, the resolution of whole-genome sequencing was critical for identifying clusters of BSI infections that would have otherwise gone unnoticed in the hospital setting. Previous reports demonstrated the utility of WGS to investigate *S. aureus* outbreaks in hospitals [6,10,14,29,34,35], though they focused on infrequent, point-source outbreaks, such as those among hospitalized infants in a neonatal intensive care unit (NICU) with an identifiable index case [29,34,35]. Also, hospital outbreak studies have often used WGS data to confirm that cases of MRSA infection were related only after initial outbreak detection by other means, including an unusual antibiogram shared between isolates [34] or the presence of an unusual strain type [10]. Collectively, these investigations identified an epidemiologically significant core genome SNP range as small as 13 SNPs [13] to as large as 40 SNPs [36] among outbreak isolates.

Our investigation confirmed that a SNP threshold under 35 was effective for detection of clustering with clear overlaps in hospital stay among adult patients in a population where MRSA transmission pathways are difficult to identify. Clusters C1, C2, C8, and C9 all had median pairwise distances between 1-25 SNPs, and patients in these clusters were diagnosed with a BSI within three months of the earliest known BSI diagnosis date in the cluster. Considering estimates of *S. aureus* neutral mutation of approximately 5-6 SNPs per genome per year, a likely scenario that explains our findings is a recent common exposure in a healthcare setting several weeks to months prior to onset of diagnosed severe disease for subjects involved in the same cluster [37]. However, isolates obtained from subjects that clustered without evidence of a hospital overlap had small ranges of SNP differences, which suggests alternative routes of MRSA transmission among individuals later diagnosed with a BSI. These might include a long-term unknown reservoir of MRSA within the hospital, or, alternatively, in the community. Based on our logistic regression, it is reasonable to investigate healthcare histories for patients at or below13 SNPs in order to find potential transmission routes. In the future WGS may be most useful to prevent infections by implementing regular screening of MRSA colonization at certain key healthcare encounters, especially for patients at high risk of BSI or another invasive MRSA infection.

It is important to evaluate different bioinformatic techniques on WGS outcomes because of the diversity in strategies across studies. Most US hospitals have not yet implemented a WGS surveillance system for infection control. Hospitals may choose to approach bioinformatic surveillance using commercial workflows with integrated processes [35], available open source options [38], or create robust in-house surveillance methods [39]. In this analysis we demonstrated that different methods of DNA sequence alignment can readily detect similar variation in SNP differences. However, isolates that would have epidemiological significance may fall outside of a typical observed SNP cut-off depending on the choice of bioinformatic tools used. We showed in our analysis that a pan-genome approach has good consistency in detecting similar SNP differences even as many strain types are included. It is likely that the bioinformatic workflow choices will continue to be variable based on user preferences and needs. However, a sliding scale [40] or a threshold range [13] could offer a more flexible alternative for including patients in transmission investigations.

Previous recommendations point to the use of a closely-related reference for alignment and phylogenetic reconstruction for identifying transmission events [15,40], and indeed this approach has been advantageous for reactive outbreak response in the past, particularly in clinical settings where MRSA infections are rare. However, *S. aureus* transmission from healthcare facilities into community settings and back into healthcare facilities demonstrates that it is important to consider the hospital and the community together as a singular reservoir of transmission [29]. Our investigation also points to the importance of long-term MRSA carriage prior to diagnosis of a BSI. An overlapping hospitalization may provide an opportunity for MRSA transmission leading to onset of asymptomatic colonization in a recipient patient, but a later infection in the recipient, such as a BSI, may not occur until weeks or months later. As a result, identification of a cluster of BSIs caused by closely related MRSA isolates is likely to occur long after the critical moment of transmission, which is an opportunity for infection control intervention. As WGS surveillance becomes more proactive for preventing spread, rather than reacting later to large clusters, it would be advantageous to implement pan-genome alignments, in place of closely related reference genomes, into workflows for WGS data analysis.

Our analysis was subject to several limitations. The long span of time between BSI onset among cluster patients and lack of an obvious transmission pathway suggests that there may have been intermediate patients who never experienced a BSI but could have carried MRSA strains that infected others. Additionally, we did not collect isolates from the hospital environment or from healthcare workers directly, so we cannot discern the role of healthcare or environmental intermediaries for transmission in the clusters.

We have provided evidence of MRSA BSI clusters among adults with a variety of prior healthcare exposures in a setting with a relatively high incidence of MRSA infections. In our study, we identified genomically tightly-related clusters where the routine epidemiological signal was weak, but with further investigation suggested healthcare exposures well before BSI presentation. Including WGS as a part of routine colonization screening that already occurs for MRSA in certain high-risk clinical settings may be helpful in identifying and preventing transmission events in areas of hospitals not regularly scrutinized by infection control staff.

## Data Availability

All data produced are available online at

https://github.com/Read-Lab-Confederation/MRSA_bloodstream_clusters

## Acknowledgements

Thank you to Laurel Glaser for assistance with the biobanking of isolates used in this study, and to Katrina Hofstetter for support in troubleshooting R code. We also thank the Penn/Children’s Hospital of Philadelphia (CHOP) Microbiome Center for performing the isolate sequencing used in this analysis.

